# Counting What Counts: Ensuring Wearable Step-Count Validity for Effective Public Health Interventions

**DOI:** 10.1101/2025.05.24.25328291

**Authors:** Bin Hu, Ahmed Abdul-Samad, Doreen Amini, Izma Ghani, Shahryar Wasif, Taylor Chomiak, Adam David

## Abstract

**Background:** Daily step counts are a widely used metric in public health and clinical practice for assessing physical activity levels, particularly in older adults and individuals with chronic conditions. However, most commercial step counters rely on forward trunk acceleration, making them prone to significant inaccuracies during vertical, non-locomotive activities such as Stepping-in-Place (SIP).

**Objective:** This study evaluated the step-count accuracy of Google Fit, a commercial accelerometer-based smartphone application, compared to Ambulosono, a wearable sensor that captures joint-specific range of motion (ROM), during music-paced SIP sessions.

**Methods:** Thirty-six participants performed multiple SIP trials using a standardized, music-based protocol. Step counts were recorded concurrently using both devices. Data were analyzed using regression modeling, k-means clustering, and Bland–Altman agreement analysis to assess accuracy, cadence responsiveness, and detection consistency.

**Results:** Google Fit consistently undercounted SIP steps by 20–60%, showing weak correlation with exercise duration (R = 0.16). Ambulosono demonstrated strong correlations with cadence (r = 0.789) and duration (R = 0.97), and uniquely captured biomechanical trade-offs such as an inverse relationship between step height and cadence. Bland–Altman analysis confirmed a systematic negative bias in Google Fit output.

**Conclusion:** These findings reveal critical limitations in commercial step counters when applied to non-forward-motion activities and highlight the advantages of ROM-based sensing for accurate and context-aware activity tracking. Ambulosono’s robust performance suggests its suitability for rehabilitation, elderly care, and home-based exercise monitoring, where step accuracy is essential for meaningful health assessment.

## Introduction

Wearable fitness technologies have become central to public health strategies and personal wellness routines, offering real-time feedback, self-monitoring, goal-setting, and motivational support across diverse populations (Evenson et al., 2015; Sullivan & Lachman, 2016). Among their many functions, daily step counts have emerged as the most universally recognized and adopted metric, serving as a benchmark for physical activity in clinical monitoring, chronic disease management, and preventive health interventions. Both healthcare professionals and patients rely on these data to assess activity levels, adherence to exercise regimens, and associated health outcomes (Saint-Maurice et al., 2020). However, the widespread assumption that commercial step counters provide uniformly accurate data across all forms of movement has increasingly been called into question.

Numerous studies have shown that commercial devices, including Google Fit, often exhibit significant discrepancies in low-speed or non-locomotive scenarios (Thorup et al., 2017; Le Masurier et al., 2004; Cleland et al., 2013). Devices that rely solely on accelerometers detect movement by measuring forward acceleration of the trunk and limbs. As a result, they can fail to register steps during activities that involve vertical movement with little or no horizontal displacement, such as stepping in place (SIP) or stationary cycling. These limitations are particularly consequential in clinical populations—such as the elderly or individuals with Parkinson’s disease—who often engage in slow, low-impact exercises as part of home-based rehabilitation or physical therapy (Hu & Chomiak, 2019).

SIP, in particular, is gaining recognition as a practical, safe, and space-efficient form of exercise. It requires minimal equipment or supervision, making it ideal for older adults, those recovering from surgery, or people with chronic conditions. Despite its clinical and public health potential, SIP poses a unique challenge for traditional step-counting algorithms. Because SIP lacks the forward locomotion typically associated with walking, the movement patterns involved—mainly vertical stepping from the hip and knee—are poorly captured by accelerometer-based systems. This biomechanical mismatch results in systematic undercounting, which can distort assessments of physical activity and impair the utility of step-count data for therapeutic or behavioral interventions.

To address these limitations, this study examines the performance of Ambulosono, a range-of-motion (ROM) based wearable sensor, in comparison with Google Fit. Ambulosono is a single-joint sensor worn above the knee that tracks hip flexion angles and gait cycle parameters through a fusion of MEMS-based gyroscopes and accelerometers (Chomiak et al., 2019). Unlike commercial devices that depend on net trunk displacement, Ambulosono focuses on the angular trajectory of the limb, allowing it to capture step-like events even when there is no forward movement. The device also includes a music-paced “GaitReminder” feature designed to standardize cadence and increase participant engagement (Hu & Chomiak, 2019).

In this study, thirty-six undergraduate participants engaged in music-based SIP sessions of variable duration, during which step data were concurrently collected by both devices. We analyzed the data using linear regression, machine learning clustering (k-means), and Bland–Altman agreement analysis. Our results show that Google Fit systematically undercounts steps during SIP by 20–60%, depending on cadence and session length, whereas Ambulosono demonstrated high correlation with step duration and cadence (R = 0.97). Moreover, Ambulosono was able to capture the biomechanical trade-off between cadence and step height, revealing meaningful insights into participant gait patterns that were completely missed by Google Fit (Figures 4–6).

**Figure 1:**
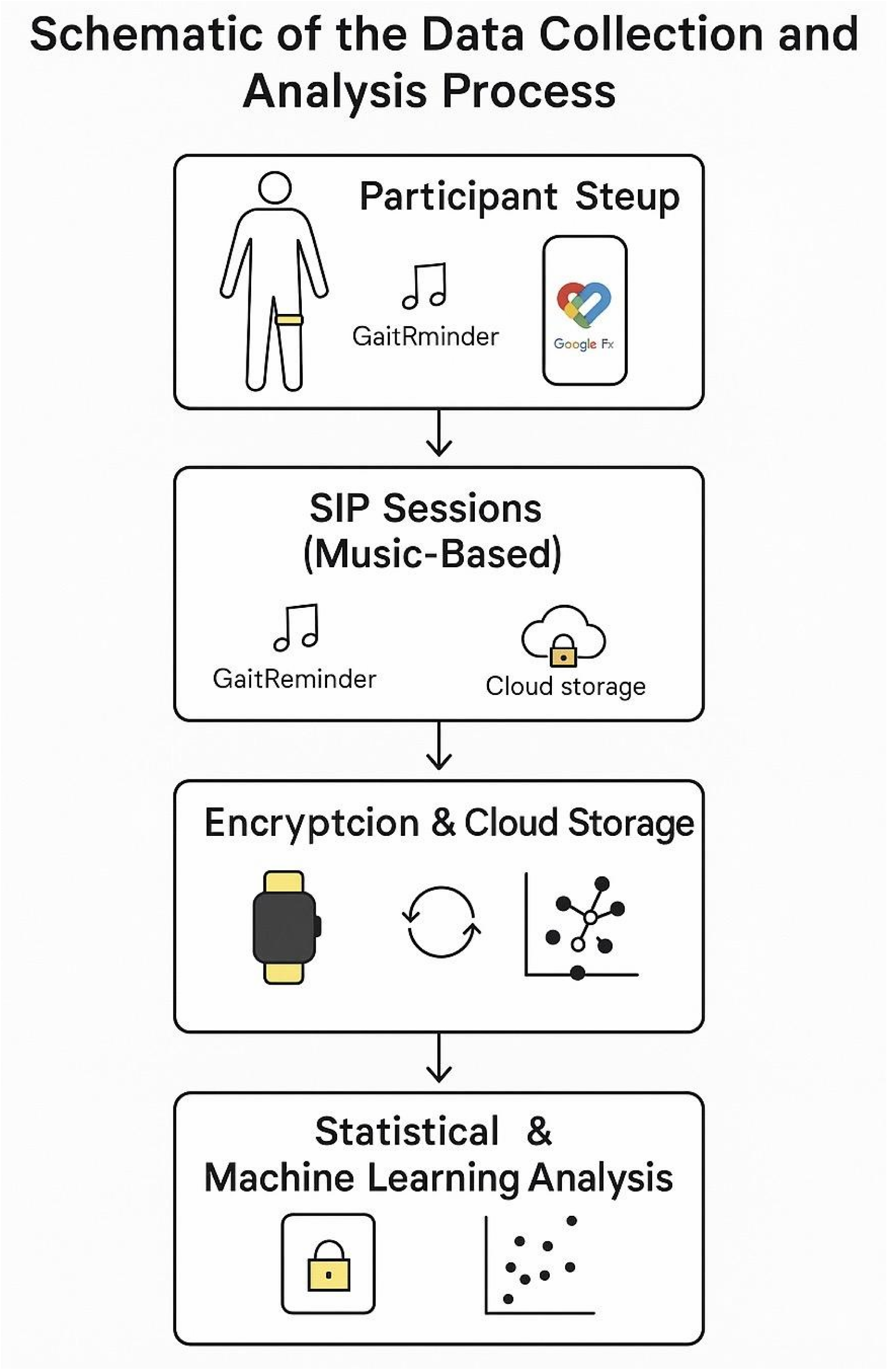
Schematic of the Data Collection and Analysis Process This figure illustrates the sequential workflow used to collect and analyze step count data during Stepping-in-Place (SIP) exercises. The process begins with participant setup, where the Ambulosono sensor is secured above the knee and the Google Fit app is activated on a smartphone. The music-based SIP sessions are managed through the Ambulosono app, which includes a GaitReminder feature to ensure consistent pacing. Step data from both devices are logged locally and encrypted before being uploaded to a secure cloud storage system. Data analysis incorporates statistical regression and machine learning clustering to evaluate step count accuracy and discrepancies between Ambulosono and Google Fit.

These findings highlight the need for context-aware, biomechanically accurate wearable devices, particularly for use in clinical and rehabilitative environments where movement patterns deviate from typical walking. The superiority of Ambulosono in this domain suggests that ROM-based tracking systems may serve as more reliable and therapeutically relevant tools for step monitoring in non-locomotive or constrained-movement activities.

## Methods

### Study Setting and SIP Protocol

This study was conducted under the auspices of the Open Digital Health (OpenDH) program at the University of Calgary—an initiative designed to expose undergraduate students to advanced research in digital health, wearable technologies, and AI-driven analytics. To evaluate wearable sensor accuracy in a realistic and engaging format, we adopted a music-based indoor Stepping-in-Place (SIP) protocol modeled after Hu & Chomiak (2019). Each participant completed multiple SIP trials, with each trial synchronized to a randomly selected music track. Songs varied in length and tempo, thereby allowing for variability in step cadence and session duration. The use of auditory pacing cues simulated real-world physical activity patterns while also minimizing performance bias and enforcing pacing regularity via Ambulosono’s built-in GaitReminder feature.

### Participants

A total of 36 undergraduate students (age range: 16–22 years) were enrolled through the OpenDH and Ambulosono training platforms at the University of Calgary. Participation was voluntary, and informed consent was obtained from all subjects in accordance with ethical standards approved by the University of Calgary Conjoint Health Research Ethics Board (CHREB). The participant cohort included individuals with varying fitness levels but excluded those with known musculoskeletal or neurological impairments. Demographic data, including age, gender, and step duration contributions, are detailed in Table 1.

**Table 1:** Demographic Information and Step Contributions by Participants.

| Participant Gender | Number of Participants | Percentage of Participants (%) | Step Duration Contributed to Dataset | Percentage of Step Duration Contributed (%) |
| --- | --- | --- | --- | --- |
| Female | 22 | 61.11 | 594 | 68.99 |
| Male | 6 | 16.77 | 125 | 14.52 |
| Prefer Not to say | 8 | 22.22 | 142 | 16.49 |
| Total | 36 | 100 | 861 | 100 |

### Devices and Data Capture

Step data were captured concurrently using two systems: Ambulosono, a single-joint wearable ROM sensor, and Google Fit, a commercially available smartphone-based step counter. The Ambulosono device integrates 3-axis MEMS-based gyroscopes and accelerometers and was affixed above the participant’s patella using an elastic thigh band (Chomiak et al., 2019). The sensor sampled motion data at 50 Hz, recording angular velocities (pitch, roll, yaw) that were subsequently used to compute gait-relevant metrics including step height, cadence, step duration, and step count. Ambulosono’s algorithm identifies step events based on hip flexion angle thresholds, enabling reliable step detection even in the absence of forward locomotion. For Google Fit measurements, a smartphone with the app pre-installed was placed in the participant’s pant pocket on the same side as the Ambulosono sensor. This placement mimicked common usage scenarios and ensured synchronized data capture. SIP sessions were manually reset between trials to isolate each recording. All data were stored locally on respective devices, then securely archived and encrypted before being uploaded to a protected cloud database.

### Music Protocol, Fatigue Assessment, and Data Analysis

Each participant stepped to a randomized playlist comprising Canadian Music Hall of Fame artists, with individual tracks spanning a range of tempos and genres. The Ambulosono app’s GaitReminder feature automatically paused playback if cadence dropped below a preset threshold, ensuring consistent movement quality across trials (Hu, 2019). Participants took 3–5 minute rest intervals between songs, during which they completed subjective fatigue and breathlessness evaluations using the Borg Breathlessness and Fatigue Rating Scales (Borg, 1998). Additionally, participants reported music preference and exercise intensity using structured questionnaires.

All step count data were analyzed using the OpenAI Advanced Data Analysis (ADA) platform, which supported both traditional statistical tools and natural language-enhanced machine learning pipelines. Linear regression was used to examine the relationship between step count and SIP duration, while k-means clustering was applied to explore device behavior across different cadence and step height profiles. Only SIP sessions with complete and synchronized data from both devices were included. Statistical significance was defined as p < 0.05. Data preprocessing included outlier filtering (step count capped at 2000 per trial), noise reduction, and quality control checks before analysis.

## Results

### Step Count vs. SIP Duration (Figure 2)

To assess how well each device tracked exercise duration, we performed linear regression analysis between SIP session duration and recorded step counts. As shown in Figure 2, the Ambulosono system exhibited a strong linear relationship between duration and step count (Steps = 281.3 × Duration + 128.7; R = 0.97), indicating high responsiveness to variations in exercise length. In contrast, Google Fit produced a notably flatter regression line (Steps = 32.9 × Duration + 407.2; R = 0.16), demonstrating minimal sensitivity to SIP duration. These results highlight Ambulosono’s capacity to accurately reflect time-dependent changes in movement, while suggesting that Google Fit’s step-counting algorithm is poorly suited for non-locomotive activity patterns.

**Figure 2:**
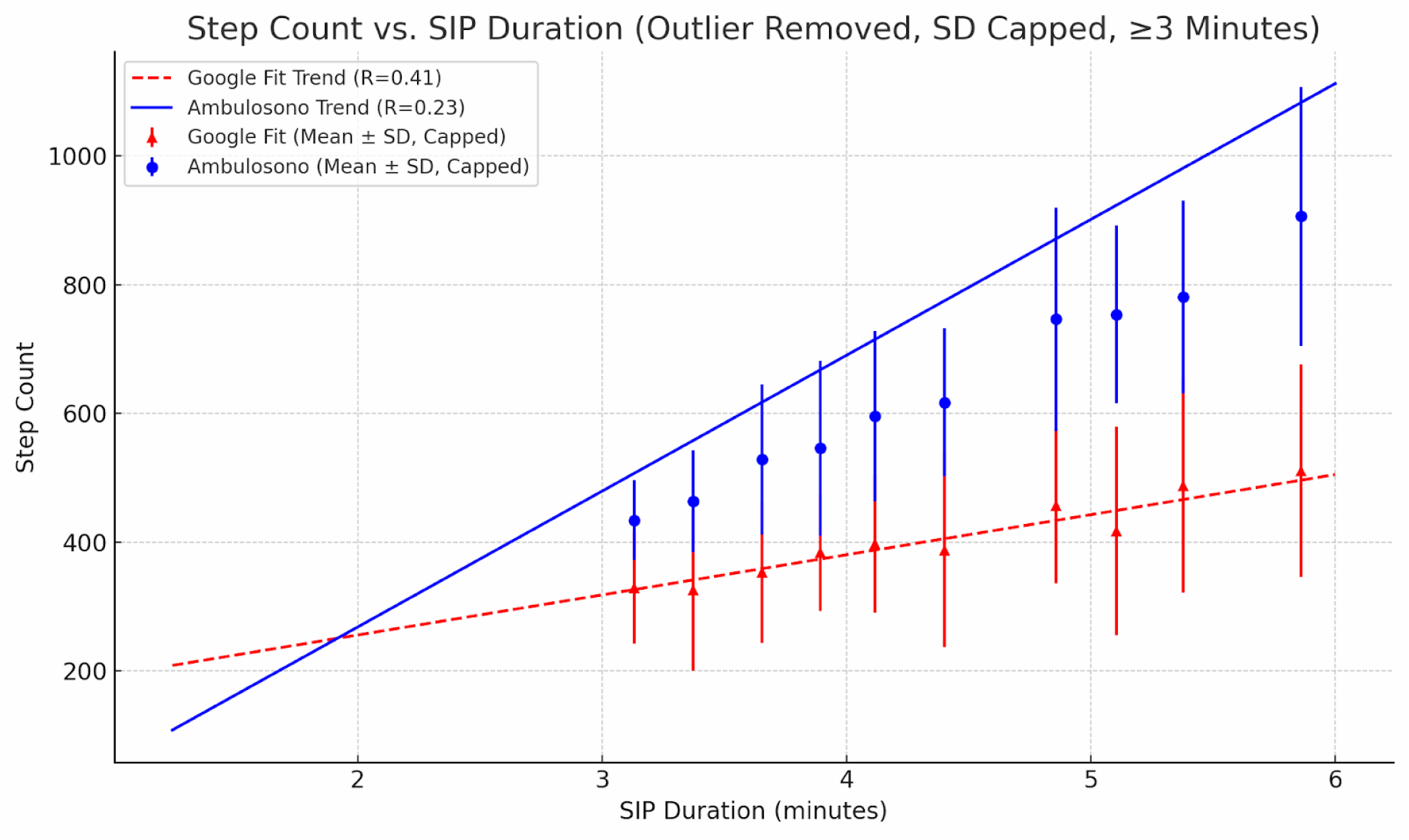
Scatter Plot Comparing Step Counts by Google Fit and Ambulosono Across Exercise Durations This figure illustrates the relationship between Stepping-in-Place (SIP) exercise duration and the step counts recorded by two wearable systems: Google Fit and Ambulosono. Data shown are restricted to SIP sessions lasting 3 minutes or longer. Each data point represents the mean step count for a duration bin of 0.25 minutes, with vertical error bars indicating the capped standard deviation (SD) to limit the influence of outliers. Blue circles denote Ambulosono mean step counts with capped SD. Red triangles denote Google Fit mean step counts with capped SD.

### Trial-Level Accuracy—Raw Step Count Trends (Figure 3A & 3B)

To explore device behavior across individual sessions, we plotted trial-level step counts for SIP durations up to 2000 steps. Figure 3A confirms that Ambulosono maintains a tight, positive linear trend across trials, reaffirming its reliability. Conversely, Figure 3B shows that Google Fit’s readings are scattered and inconsistent, with frequent plateaus and nonlinear deviations that do not correspond with increased session duration. This reinforces the earlier finding that Google Fit lacks the granularity required to detect vertical stepping with consistency.

**Figure 3A and 3B.**
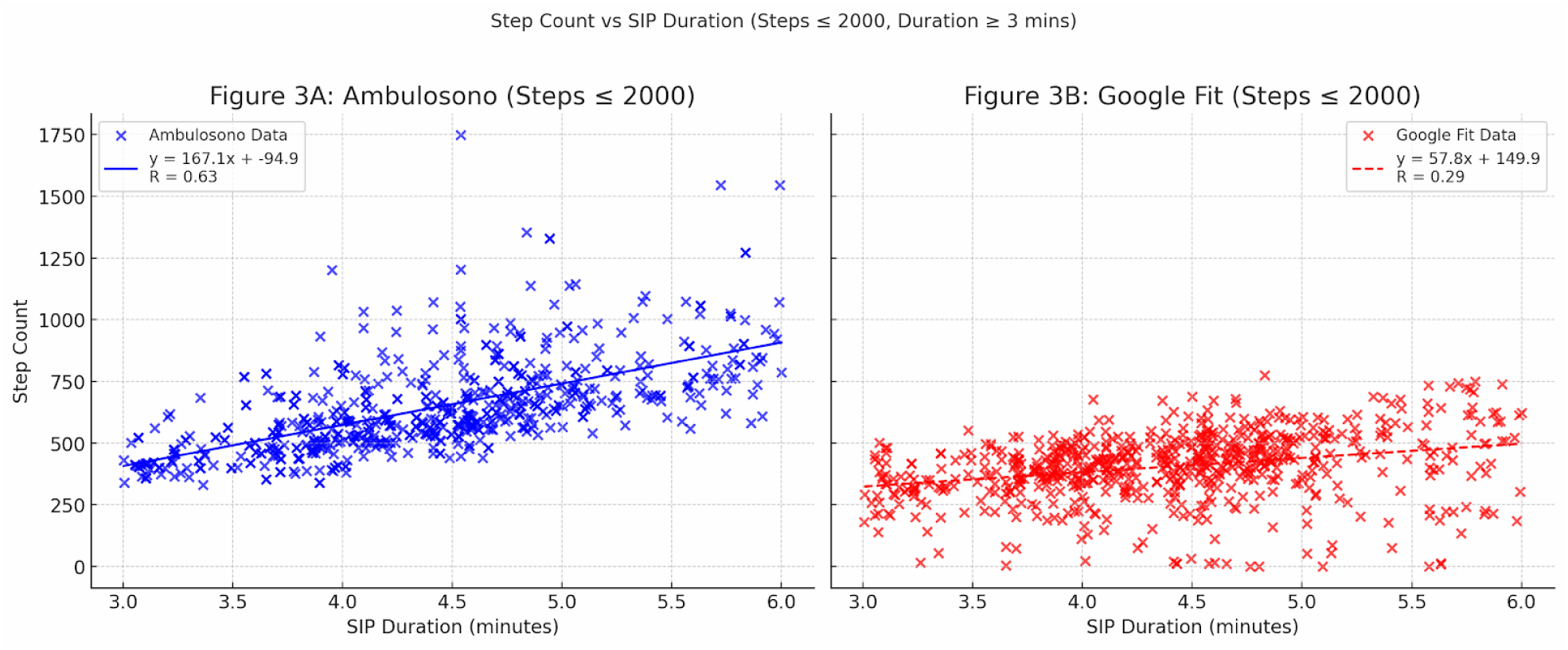
Step Count vs. SIP Duration for Ambulosono and Google Fit (Filtered to Steps ≤ 2000) These side-by-side scatter plots illustrate the linear relationship between Stepping-in-Place (SIP) duration (in minutes) and step counts measured by two wearable tracking systems: Ambulosono (Figure 3A) and Google Fit (Figure 3B). Unlike Figure 2—which displayed binned means with error bars—this figure presents raw session-level data for each device, with all sessions capped to step counts ≤ 2000 to remove outlier-driven distortions. Each figure includes individual SIP trials (dots) with regression line and equation.

### Step Height vs. Step Count Correlation (Figure 4A & 4B)

**Figure 4A-B.**
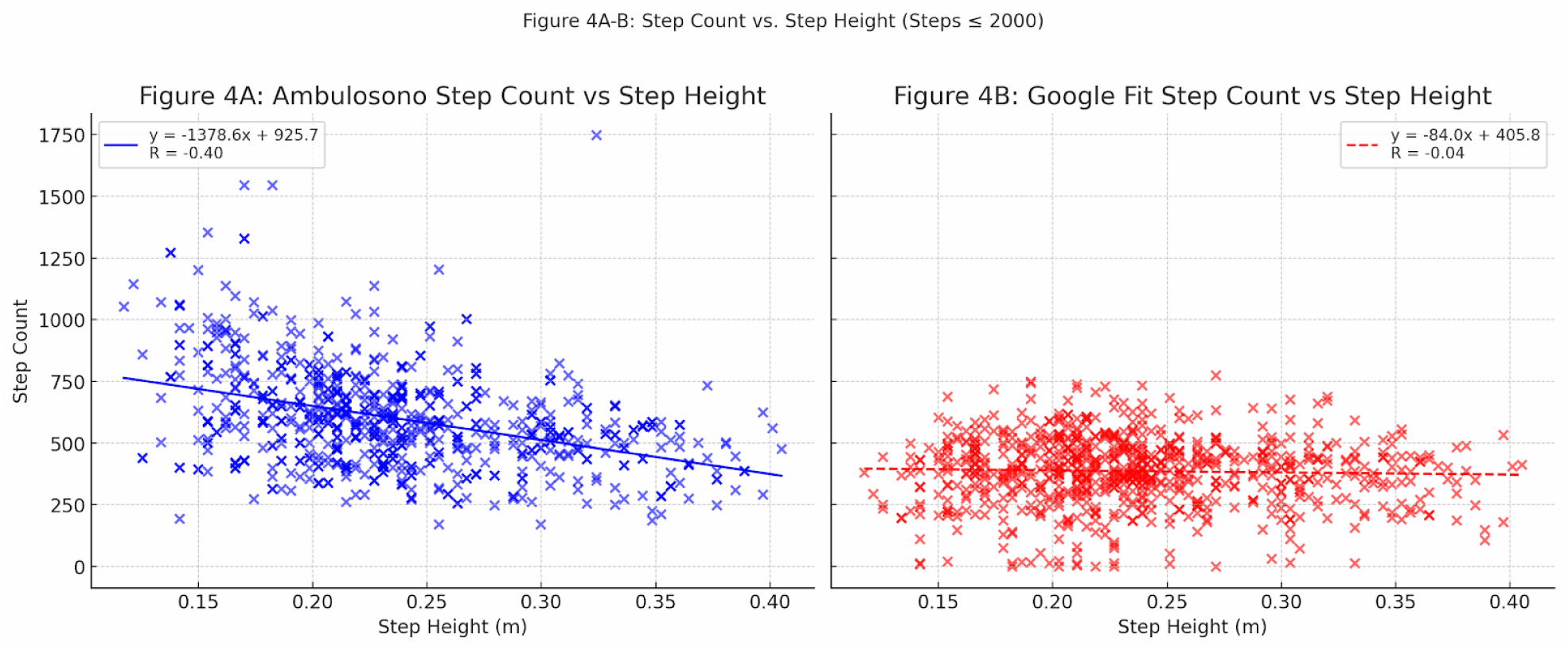
Relationship Between Step Height and Step Count for Ambulosono and Google Fit These side-by-side scatter plots depict the relationship between step height (in meters) and step count for Ambulosono (Figure 4A) and Google Fit (Figure 4B). Only SIP sessions with total step counts ≤2000 were included to ensure clean visualization. Each plot includes a regression line with its corresponding equation and R-value. Ambulosono data are shown in blue with a solid trendline, while Google Fit data are shown in red with a dashed line. Both axes are scaled consistently to allow direct comparison of trend behavior between the two systems.

Next, we examined whether step height influenced the devices’ ability to detect steps. As depicted in Figure 4A, Ambulosono showed a moderate negative correlation between step height and step count (R = -0.37), suggesting that higher steps may reduce cadence, thus lowering total step counts in a given time frame. In contrast, Figure 4B shows that Google Fit had no meaningful correlation (R = -0.07) between these variables. These results challenge the assumption that step height is a limiting factor in accelerometer-based trackers; rather, the ability to detect rhythmic cadence appears to be more critical.

### Cadence vs. Step Count Correlation (Figure 5A & 5B)

Cadence was strongly correlated with total step counts in both systems, but Ambulosono again outperformed Google Fit. As shown in Figure 5A, Ambulosono had a correlation of r = 0.789, while Google Fit achieved r = 0.729 (Figure 5B). These results indicate that as participants increased their step rate, both devices registered more steps—but Ambulosono’s tighter relationship suggests higher sensitivity to pacing variations. This reinforces the premise that cadence is a more reliable predictor of step count accuracy than step height in SIP scenarios.

**Figure 5A-B.**
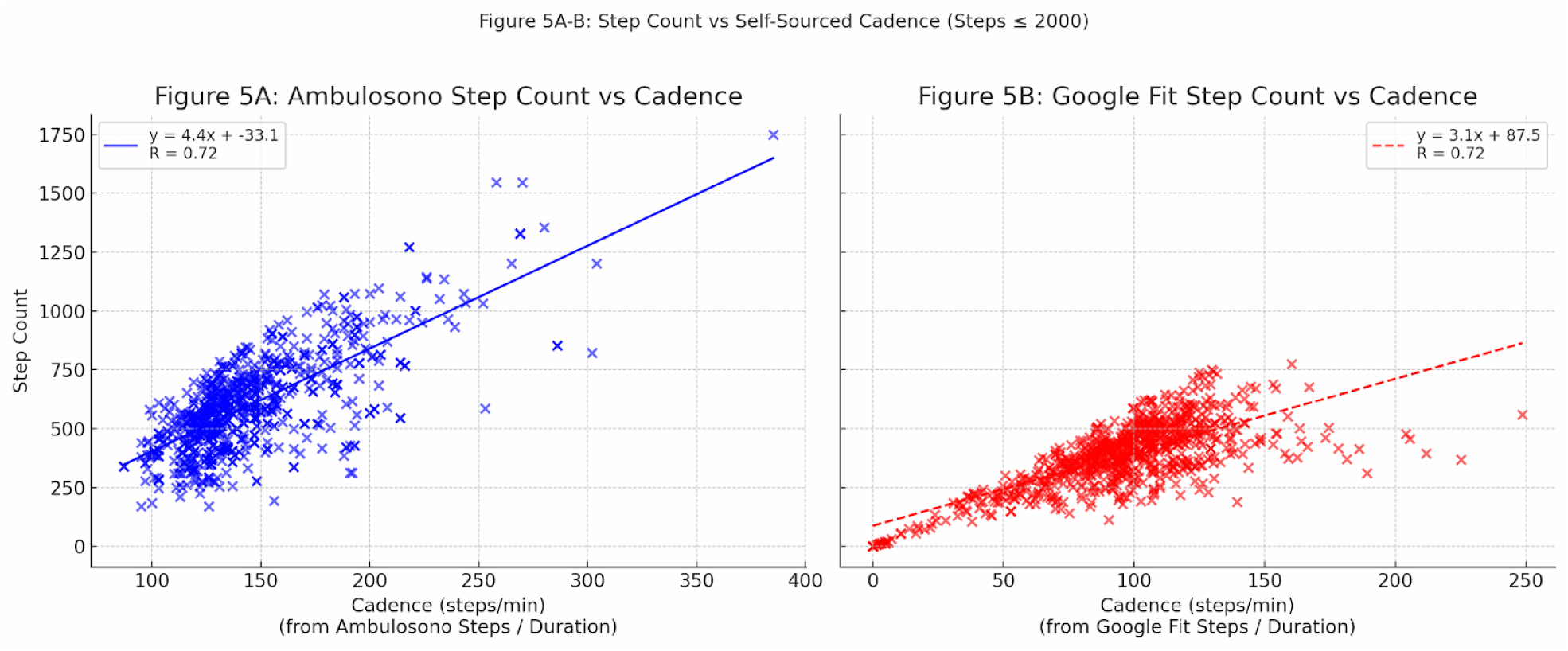
Step Count vs. Device-Sourced Cadence During Stepping-in-Place These scatter plots illustrate the relationship between cadence and step count for Ambulosono (5A) and Google Fit (5B), using cadence values calculated independently for each device by dividing total step count by trial duration (in minutes). Only trials with ≤2000 total steps were included. Regression lines with equations and R-values are overlaid, using blue for Ambulosono and red (dashed) for Google Fit. Axes are scaled consistently to enable direct comparison of device behavior across self-referenced cadence metrics.

### Cadence vs. Step Height – Quantile Regression (Figure 6A & 6B)

**Figure 6.**
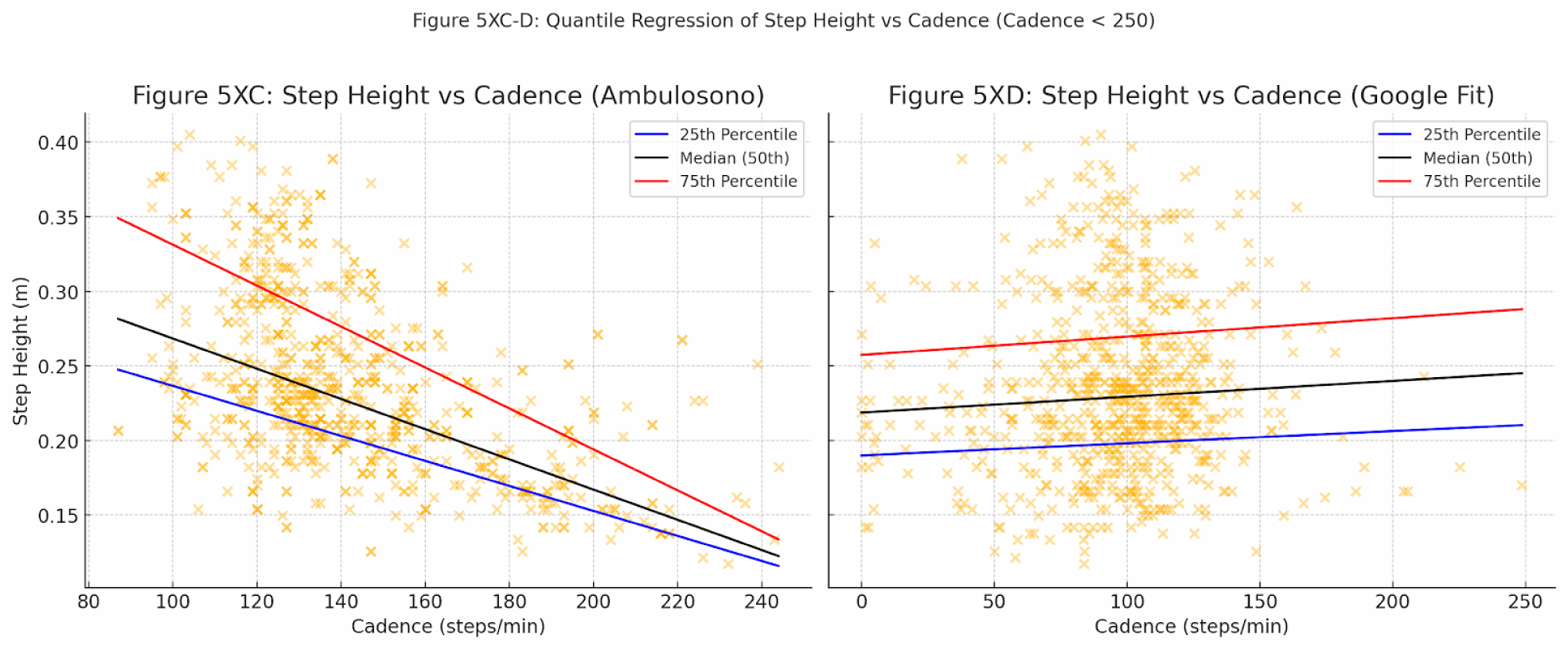
Quantile Regression of Step Height vs Cadence for Ambulosono and Google Fit These side-by-side quantile regression plots assess how cadence predicts step height across the 25th, 50th (median), and 75th percentiles. Ambulosono data are shown in Figure 6A using internal cadence and height measurements. Google Fit data are shown in Figure 6B, with cadence derived from Google Fit step count and duration, and step height sourced externally from Ambulosono. Colored lines represent the quantile regression trends: blue for the 25th percentile, black for the median, and red for the 75th percentile.

To evaluate biomechanical patterns, we performed quantile regression on the relationship between cadence and step height. Figure 6A shows that Ambulosono captured a clear inverse relationship across all quantiles: higher cadence was associated with lower step height, suggesting a physiological trade-off. The quantile slopes varied, implying individual differences in how users balance rhythm and amplitude. In contrast, Google Fit-derived cadence (Figure 6B) displayed no structured trend, highlighting the system’s inability to track vertical displacement or resolve participant-specific gait patterns.

Discrepancy Analysis & Agreement Testing (Figures 7A, 7B, and 8) Step Count Discrepancy (Figure 7A & 7B)

**Figure 7.**
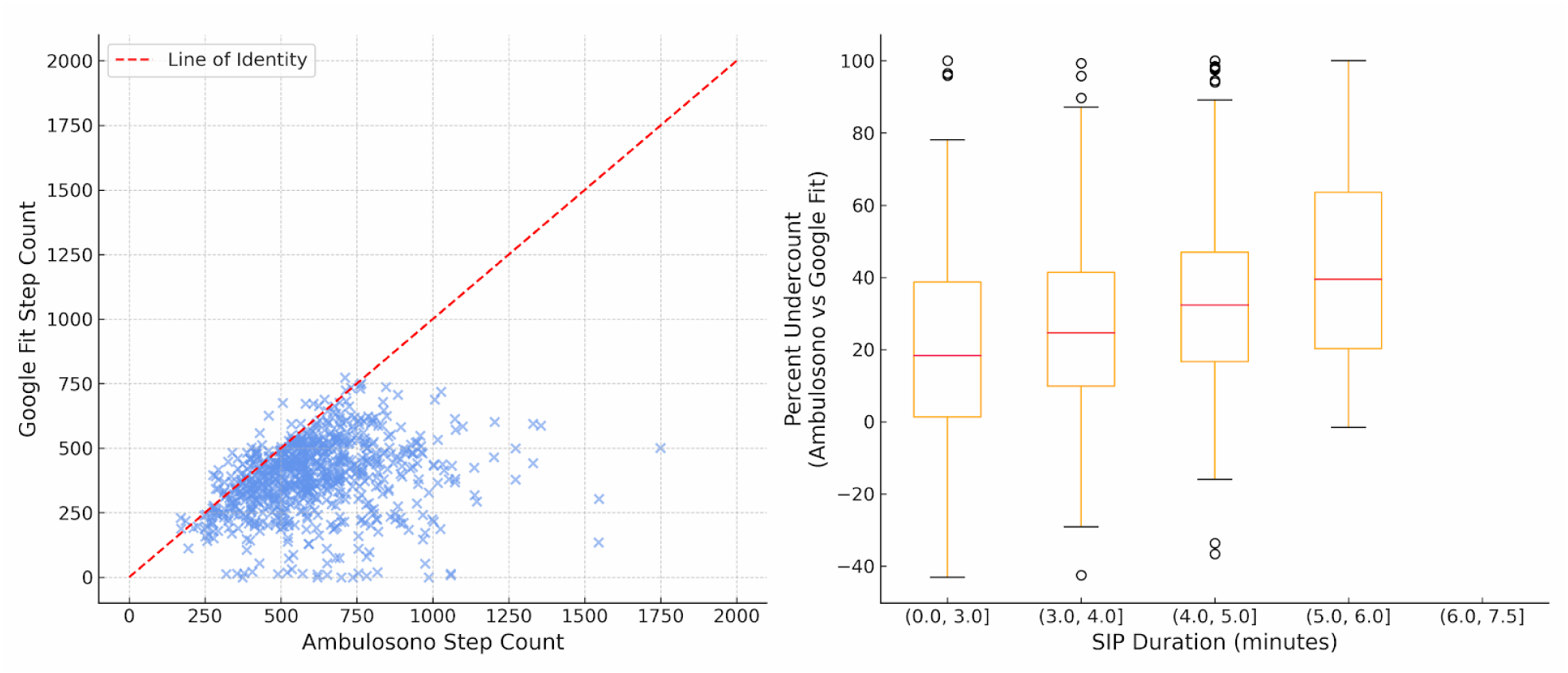
shows the step count discrepancies between Ambulosono and Google Fit. The left (A) scatter plot compares raw step counts recorded by Google Fit and Ambulosono during SIP trials (≤2000 steps). Each point represents a single trial. The red dashed line is the line of identity (y = x), indicating perfect agreement. Points below the line represent trials in which Google Fit undercounted steps relative to Ambulosono. The right panel (B) is the box plot showing the distribution of percentage discrepancy between the two systems (expressed as a percent of Ambulosono count) grouped by SIP duration bins. Each box summarizes the median, interquartile range, and outliers of discrepancy for each duration range.

Scatter plot analysis (Figure 7A) reveals that nearly all trials fell below the line of identity, confirming that Google Fit consistently undercounts steps relative to Ambulosono. Discrepancies were not limited to specific trial durations, as seen in the box plot (Figure 7B), where median undercounts ranged from 30% to 60% across all SIP bins. This undercounting remained systematic and duration-independent, suggesting that Google Fit does not recalibrate over longer sessions.

### Bland–Altman Agreement (Figure 8)

The Bland–Altman plot shows a consistent negative bias, with Google Fit underestimating step counts by an average margin that remained stable across step ranges. The broad limits of agreement indicate substantial trial-to-trial variability, reaffirming that the two systems are not interchangeable in SIP contexts. The findings collectively support the hypothesis that ROM- based devices provide more stable and sensitive measurement in non-locomotive exercise modalities.

**Figure 8.**
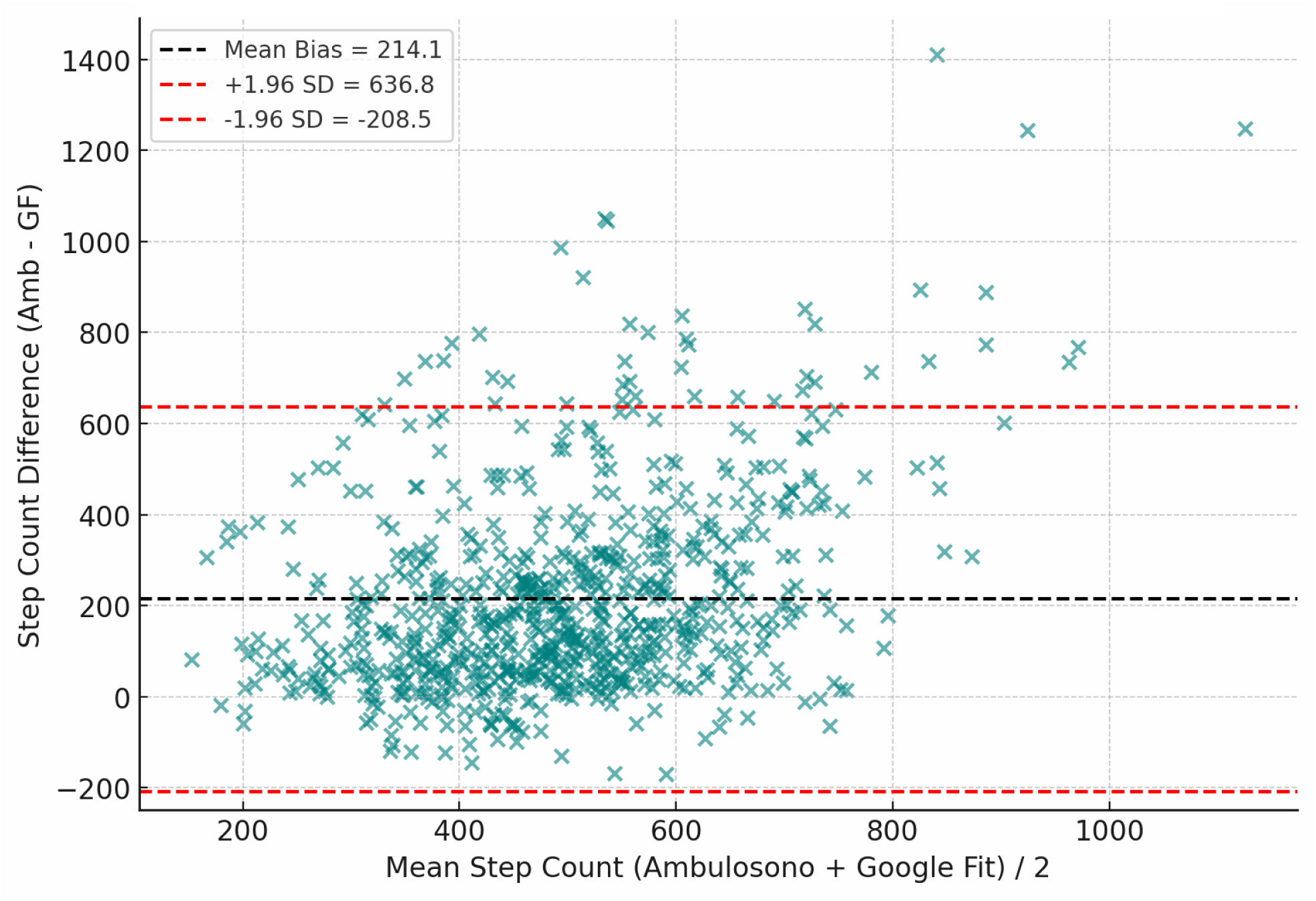
Bland–Altman Plot of Step Count Agreement Between Ambulosono and Google Fit This plot illustrates the agreement between Ambulosono and Google Fit step counts using the Bland–Altman method. Each point represents one SIP trial. The X-axis displays the average step count across both devices, while the Y-axis shows the difference (Ambulosono – Google Fit). The solid black line represents the mean bias, while the dashed red lines denote the limits of agreement (mean ± 1.96 × SD of the difference). These bounds define the range within which 95% of differences are expected to fall.

All the above results are summarized in Table 2.

**Table 2:** Summary of key analysis and results.

| Key Insight | Device Findings | Significance |
| --- | --- | --- |
| Regression (Steps vs. Duration) | Ambulosono: R = 0.97; Google Fit: R = 0.16 | Strong contrast in time-sensitivity |
| Raw Trial Trends | Ambulosono: Consistent; Google Fit: Scattered | Trial-level evidence of poor tracking by Google Fit |
| Step Height Influence | Ambulosono: Moderate inverse; Google Fit: None | Step height not main limiting factor |
| Cadence vs. Steps | Ambulosono (r = 0.789) > Google Fit (r = 0.729) | Cadence is the key determinant |
| Cadence-Step Height Trade-off | Captured only by Ambulosono | Reflects authentic gait properties |
| Discrepancy +<br>Bland–Altman | Systematic<br>undercounting by<br>Google Fit | Devices are not<br>interchangeable |

## Discussion

This study provides critical insights into the limitations of commercial step-counting technologies—particularly Google Fit—when used during stationary exercises such as Stepping-in-Place (SIP). Our findings clearly demonstrate that Google Fit systematically undercounts steps by 20–60%, an error margin that persisted across session durations and cadence levels.

These discrepancies, also reported in prior studies involving other consumer-grade wearables (Thorup et al., 2017), raise significant concerns regarding the generalizability and clinical reliability of step data generated by accelerometer-based fitness trackers in non-locomotive contexts.

The observed discrepancies stem from fundamental differences in device architecture and sensing strategies. Google Fit’s reliance on accelerometers tuned for forward trunk motion makes it insensitive to the vertical and joint-specific dynamics of SIP, where there is minimal translational movement. In contrast, the Ambulosono system, which captures joint-specific angular motion, was shown to track step counts with high fidelity across trials, durations, and pacing conditions. The system’s strong correlation with cadence (r = 0.789) and SIP duration (R = 0.97) highlights the robustness and precision of ROM-based monitoring platforms in capturing exercise behavior beyond traditional walking. (see also Sullivan et al., 2016; Rodriguez et al., 2018; Cristia et al., 2020; Li et al., 2015; Chomiak et al., 2017; Iovanel et al., 2023; An et al., 2019; Yang et al., 2010; Retscher et al., 2020; Douma et al., 2018; Case et al., 2015).

An important biomechanical insight revealed by Ambulosono’s dataset is the inverse relationship between cadence and step height. As cadence increases, participants tend to adopt lower step heights, reflecting an energy-conservation trade-off consistent with overground gait dynamics.

This relationship was clearly captured in quantile regression plots, with varying slopes across user profiles, suggesting participant-specific strategies in step execution. Google Fit, by contrast, was unable to detect these nuanced variations, reinforcing its limited utility for biomechanical or rehabilitative assessments. Furthermore, Bland–Altman analysis confirmed a stable negative bias in Google Fit step estimates, suggesting that its algorithmic limitations are systematic rather than random and unlikely to be resolved through longer usage or repetition. (see also Chen et al., 2018; Charlton et al., 2022; Ngueleu et al., 2019; Scherrenberg et al., 2022; Le et al., 2004; Chomiak et al., 2015; Evenson et al., 2015; Bonomi et al., 2018; Kwapisz et al., 2011; Cleland et al., 2013).

These findings underscore an important methodological point: cadence—not step height—is the more reliable predictor of step count in SIP activities. While it has been assumed that higher step heights could challenge accelerometer sensitivity, our data show that cadence drives sensor responsiveness more directly. This suggests that future commercial algorithms should prioritize pacing and rhythm detection over amplitude-based thresholds when adapting step counters for non-forward activities.

From an applied standpoint, these results have practical and clinical implications. SIP is increasingly promoted for older adults, individuals recovering from surgery, or those managing chronic illness, due to its low spatial demand, safety, and accessibility (Tudor–Locke & Bassett, 2004; Hu & Chomiak, 2019). In such contexts, accurate monitoring is essential for tracking therapeutic progress, compliance, and health outcomes. ROM-based devices like Ambulosono can not only quantify SIP steps with precision, but also provide insights into motion quality, fatigue, and adaptation patterns, enabling a richer understanding of patient mobility.

Our findings also point to new directions for wearable device development. Algorithms must be tailored to account for contextual variables, including movement direction, user-specific cadence thresholds, and non-traditional gait patterns. As shown by Rodriguez et al. (2018) and Scherrenberg et al. (2022), environmental and algorithmic variables profoundly affect measurement precision. The development of context-aware sensing systems, supported by real-time feedback mechanisms like GaitReminder, could dramatically improve data validity in clinical and home-based exercise tracking.

Finally, we acknowledge several limitations in our study. The participant pool was restricted to young, healthy university students, which may limit the generalizability of findings to older or more heterogeneous populations. While our SIP protocol was standardized and musically guided, real-world adherence and environmental variability could introduce additional noise not accounted for here. Despite these constraints, our study offers compelling evidence that ROM- based sensing systems can outperform traditional accelerometer-based step counters in constrained, vertical-movement exercises, and should be prioritized in future wearable design and public health monitoring tools.

## Conclusion

This study evaluated the accuracy of two wearable systems—Google Fit and Ambulosono—during Stepping-in-Place (SIP), a rehabilitation-focused activity characterized by vertical, non-forward motion. Results revealed that Google Fit consistently undercounted steps by 20–60%, failing to track SIP accurately due to its reliance on forward trunk movement. In contrast, Ambulosono, a ROM-based wearable, demonstrated high sensitivity to cadence and exercise duration, capturing step patterns with significantly greater accuracy.

Ambulosono also detected biomechanical trade-offs, such as the inverse relationship between step height and cadence, which Google Fit could not resolve. These findings underscore the limitations of commercial accelerometer-based trackers in constrained-motion contexts and highlight the need for context-aware, ROM-based solutions in clinical and home-based care. As wearable technologies become increasingly central to health monitoring, algorithmic refinement and motion-adaptive design will be critical to ensuring data accuracy and clinical utility.

## Data Availability

All data produced in the present study are available upon reasonable request to the authors

## Acknowledgments

We would like to extend our sincere gratitude to Adam David for his role in organizing the research collection and serving as a progenitor for this study. Our thanks also go to Shahryar Wasif, Farhan Raza, and Elbert Tom for their role in advising and assisting with statistics and stylistic choices. We express our appreciation to the OpenDH Program at the University of Calgary, and specifically to Thowfeeq, Zuha, Victor, Doreen, and Izma for their assistance and support in data collection. Lastly, our heartfelt thanks to all the University of Calgary students, their families, and friends who participated in our study. Your involvement was fundamental to the success of our research.

## Notes

### Competing Interest Statement

The authors have declared no competing interest.

### Funding Statement

This work was partially funded by Alberta Ministry of Mental Health

### Author Declarations

A total of 36 undergraduate students (age range: 16 to 22 years) were enrolled through the OpenDH and Ambulosono training platforms at the University of Calgary. Participation was voluntary, and informed consent was obtained from all subjects in accordance with ethical standards approved by the University of Calgary Conjoint Health Research Ethics Board (CHREB).

